# AI-assisted nurse-led skin cancer screening in a teledermoscopy framework: a multi-site evaluation with one million lesions

**DOI:** 10.64898/2026.10.01.26364543

**Authors:** Noor E Karishma Shaik, Nandakishor Desai, Kyle Wang, Lara Wild, Adam G. Dunn, Amanda Oakley, Marimuthu Palaniswami, Johan Vendrig

## Abstract

Nurse-led skin cancer screening extends specialist reach but depends on the nurse selecting which lesions to forward for diagnosis, and performance varies with experience. We evaluated whether real-time decision support using artificial intelligence (AI) improves malignancy detection in routine nurse-led teledermoscopy screening across MoleMap clinics in New Zealand and Australia (January 2024–July 2025). In this real-world study, clinics using AI decision support were compared with standard clinics. Nurses examined patients, captured dermoscopic images, and forwarded selected lesions to teledermatologists, who provided the reference diagnosis. The analytic cohort comprised 1,102,382 lesions from 98,422 patients across 577 sites. AI-assisted screening was associated with a higher malignancy detection rate than standard screening (25.5 vs 15.7 malignancies per 1,000 lesions; odds ratio adjusted for nurse experience 1.73, 95% CI 1.68–1.77), a finding consistent across all nurse-experience tiers and the three major malignant subtypes. AI assistance was also associated with a shift in recommended management: 21 additional intervention recommendations and 13 additional safety-netting recommendations (self-monitoring, short-term follow-up, or specialist referral) per 1,000 lesions, approximately balanced by 34 fewer no-action recommendations. The reference standard was teledermatologist diagnosis and allocation was not randomised; findings are therefore associational and require prospective, outcome-based confirmation.

## INTRODUCTION

Skin cancer is among the most common cancers globally and a leading cause of preventable mortality in high-risk phenotypes. In New Zealand, melanoma carries one of the highest age-standardised incidences globally, approximately 40.5 per 100,000 persons per year in 2023 with mortality exceeding 3 per 100,000 [1–3]. Prognosis is strongly dependent on tumour thickness at diagnosis: five-year survival exceeds 95% for melanomas detected below 1 mm Breslow thickness but falls below 50% for lesions greater than 4 mm [4]. Early detection is therefore critical to mortality reduction. However, population-level access to expert dermatological assessment is unevenly distributed, with rural, remote, and underserved populations experiencing longer diagnostic delays and reduced access to specialist evaluation. These delays are associated with presentation at more advanced stages and worse survival outcomes [5].

Nurse-led skin cancer screening models have been increasingly adopted to address these access constraints. In such models, trained nurses acquire demographic, medical and lesion metadata as well as total-body photography, close-up and dermoscopic images at the clinics. Selected images are then transmitted for asynchronous assessment by teledermatologists, who provide diagnosis (the diagnostic reference for this study) and recommend subsequent clinical management [6,7]. The value of these models rests on a thorough skin examination: the nurse decides which lesions warrant image capture, and which are forwarded for specialist opinion. This workflow scales specialist judgement across geographically distributed clinics, but it places substantial demands on image acquisition and lesion selection, and on the dermatologist’s ability to discriminate malignant from benign lesions from a high-volume image stream. Nurse experience affects which lesions are recognised as warranting image capture and forwarding, and how thoroughly each anatomical region is documented [8]. Per-lesion teledermatologist assessment time is also variable, reflecting experience, training, confidence, and lesion and patient factors. High-volume image streams may increase diagnostic error risk, particularly for subtle lesions [9,10]. These sources of variability, particularly inconsistent lesion selection by nurses of differing experience create opportunities for decision support systems to standardise performance and improve diagnostic yield.

Clinical decision support with artificial intelligence (AI) has been proposed as a means of augmenting lesion assessment within such workflows by providing consistent, image-based risk estimates at the point of care. Deep learning approaches applied to dermoscopic images have demonstrated high diagnostic performance in retrospective studies using curated datasets, with systematic reviews reporting performance comparable to expert dermatologists under controlled conditions [11–14]. Most prior evaluations of AI in dermatology have focused on algorithmic performance under retrospective or experimental conditions, benchmarking AI against dermatologists on curated image datasets [15–17]. They documented high diagnostic accuracy in these settings but have consistently identified a gap in evidence for real-world clinical impact and workflow integration. Evidence from real clinical settings remains scarce. One systematic review screened 760 studies and found only 18 that developed or validated skin-cancer AI in a clinical environment [18]. This scarcity reflects the challenges of implementing AI in routine clinical workflows, where factors such as image quality variability, diverse patient populations and integration with existing clinical processes differ substantially from controlled research environments.

More recent work has evaluated AI as an assistive tool in clinical practice with real-world deployments emphasising the reduction of unnecessary specialist referral of benign lesions as a principal benefit [19]. Relative to head-to-head accuracy studies benchmarking AI against dermatologists, our effect sizes are not directly comparable. The comparator is an established specialist teledermatology service, and AI acted only as decision support at the nurse stage, not as an autonomous diagnostic tool. While an international study on the same platform found dermatologist agreement sufficient to support teledermoscopic diagnosis (κ 0.81– 0.97 for melanoma) [20], teledermoscopic triage safely streams referred lesions with acceptable diagnostic accuracy [21], and sequential digital dermoscopy detects thinner melanomas over time, demonstrating the clinical utility of teledermoscopic monitoring [22], the teledermatologist reference standard is not infallible. Any residual misclassification by teledermatologists applies equally to both arms, so it is unlikely to bias the comparison between AI-assisted and standard screening.

This study evaluates whether real-time AI decision support improves malignancy detection when integrated into routine nurse-led screening. Secondary aims were to assess whether improvements varied according to nurse experience and to examine detection across major malignant lesion subtypes. We also characterised the effect of AI on clinical action recommendations and evaluated the appropriateness of biopsy and excision recommendations under each pathway. Throughout the study, clinicians retained full responsibility for diagnosis and clinical management. AI served as decision support only. By evaluating AI within an established nurse-led screening model, this study addresses whether decision support can improve malignancy detection without substantially increasing unnecessary procedures, thereby enhancing existing clinical workflows.

## METHODS

### Study Design and Setting

This was a prospective, multi-site real-world study of AI decision support in routine nurse-led teledermoscopy screening within the MoleMap network in New Zealand and Australia. Screening visits between 1 January 2024 and 31 July 2025 were included. Two care pathways operated within the network: standard nurse-led screening and AI-assisted screening. A clinic’s pathway was determined by whether the AI system had been deployed at that site. Allocation was at the clinic level and was not randomised. AI decision support was already in routine use at participating New Zealand clinics before the study began, with adoption rates of 69.8% of visits in January 2024 increasing to 78% by July 2025. The comparison therefore reflects natural variation in clinic-level AI deployment rather than a controlled rollout or intervention trial. The study was approved by the University of Melbourne Human Research Ethics Committee (HREC ID 30352). It used de-identified data from patients who had consented to research use of their information and images under MoleMap’s standard data-use agreement.

### Participants and data

Data were derived from the MoleMap clinical database. Eligible visits were full-body skin screening visits (Full Body MoleMap, SkinCheck, or SkinCheck+) for new and returning patients. Single-lesion assessments and ancillary services were excluded, as these did not constitute full-body screening. All eligible visits during the study period were included. No formal sample-size calculation was performed. Australian visits contributed to the standard-care cohort only. AI-assisted screening had been introduced at just two Australian sites in the final two months of the study, but too few visits for meaningful comparison. The unit of observation was the lesion. Each lesion represents a distinct skin site, imaged with one or more clinical and dermoscopic photographs and assigned a diagnosis by a teledermatologist. Patients contributed a mean of 9.9 (median 3) lesions per visit, and follow-up patients could have the same lesion re-imaged at later visits, so lesion assessments are clustered within visits, patients, nurses, and clinics. Lesion- and patient-level fields included age, patient-estimated Fitzpatrick phototype, and nurse experience. Nurses were grouped by time since initial dermoscopy training into three tiers: 0–2 years (Tier I, novice), 2–4 years (Tier II, intermediate), and >4 years (Tier III, experienced). Patient sex was recorded for approximately 78% of visits. Lesion anatomical site and complete patient sex were not included in the study data extract and could not be analysed. The denominator for all rates was lesions with a completed teledermatologist diagnosis. A count of AI-assessed but undiagnosed lesions existed for AI-assisted visits only and was excluded to keep the denominator comparable across arms. Full lesion accounting is given in the Results Section and **Figure 1**.

**Figure 1.**
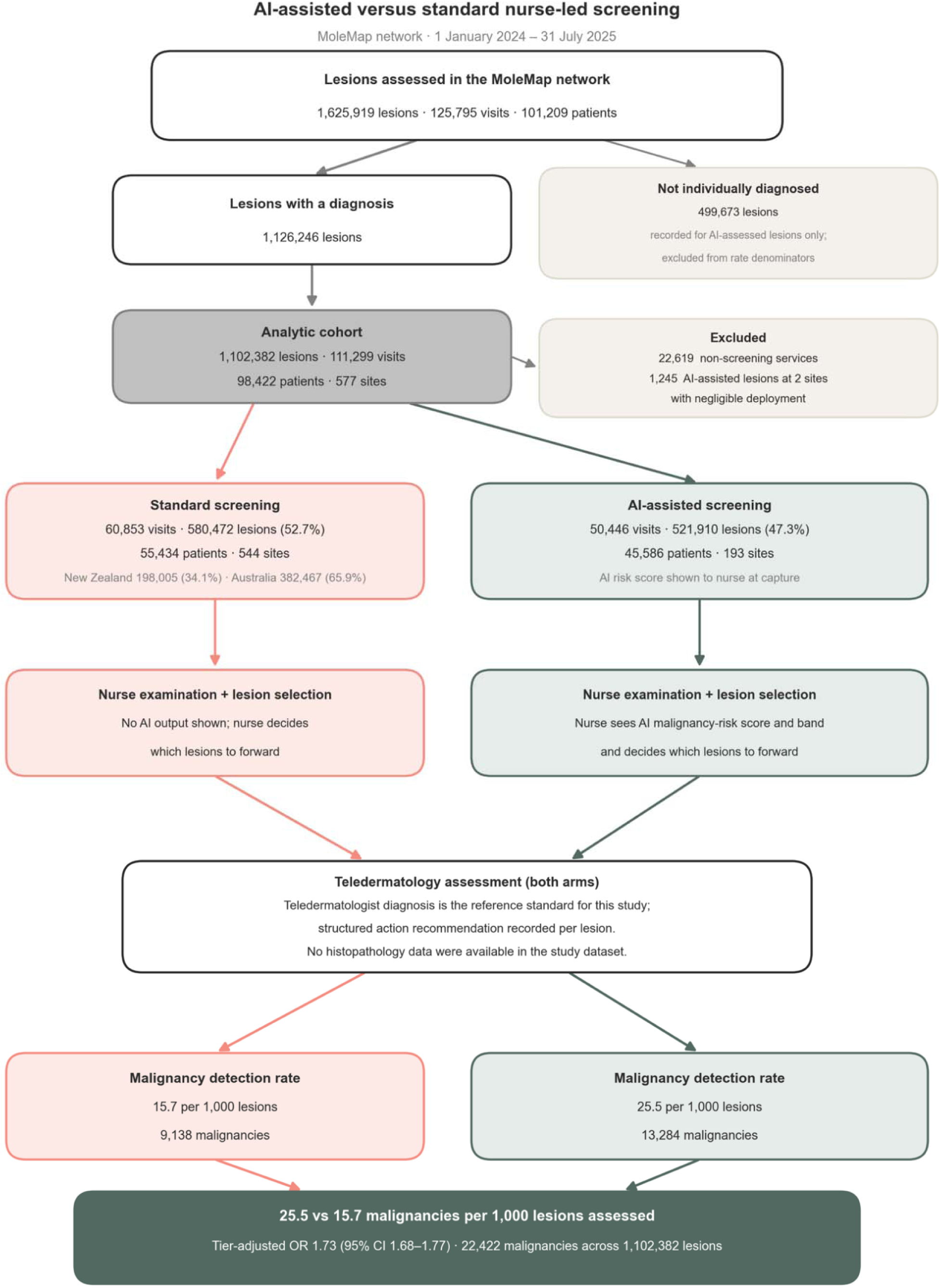
Study flow: AI-assisted versus standard nurse-led screening. Schematic representation of two-arm study flow with full lesion accounting. Colours denote screening pathway: green, AI-assisted screening; coral, standard screening. Of 1,625,919 lesions assessed across the network, 1,126,246 had a completed teledermatologist diagnosis; 499,673 lesions received an AI assessment without an individual diagnostic record and, because this information is recorded for AI-assessed lesions only, are excluded from rate denominators. After excluding non-screening services and Australian AI-assisted visits, the analytic cohort comprised 1,102,382 lesions. The nurse saw an AI malignancy-risk score and band and could use it when deciding which lesions to forward in AI-assisted clinics. No AI output was shown in standard clinics. Nurse-selected lesions from both arms were sent for teledermatology assessment, which produced the reference diagnosis and an action recommendation. No histopathology data were available in the study dataset. OR, odds ratio; CI, confidence interval.

### Reference standard and outcomes

The teledermatologist diagnosis was the reference standard for every lesion. Histopathology results were not available in the study data. Where a lesion was recommended for biopsy or excision, the procedure was performed outside the imaging pathway and neither the procedure nor its result was recorded in the dataset. Diagnoses used the MoleMap taxonomy (Diagnose Group, Major Category, Minor Category). Malignant lesions were classified as melanoma, basal cell carcinoma, squamous cell carcinoma, or other malignancy. The primary outcome was the malignancy detection rate: teledermatologist-diagnosed malignant lesions per 1,000 lesions assessed. Secondary outcomes were the detection rate for each major malignant subtype and the distribution of recommended clinical actions. Recommended actions were grouped into three categories: no action, safety-netting (self-monitoring, short-term follow-up, or referral to a GP or specialist), and intervention (biopsy, excision, or superficial treatment). We also report the rate of biopsy or excision recommendations and the number of such recommendations per diagnosed malignancy. These represent teledermatologist recommendations, not confirmed procedures or histopathology results.

### AI as Decision Support

The AI system was developed on a separate archive of MoleMap dermoscopic lesions with no overlap at the patient or lesion level with the visits analysed in this study. It comprised two models operating in parallel: a primary lesion-triage model, and a secondary disambiguation model optimised to distinguish actinic keratosis (AK) from squamous cell carcinoma in situ (SCCiS), a recognised area of dermoscopic ambiguity. Both shared a hybrid backbone combining a convolutional neural network for local texture and morphological features with a vision-transformer module capturing global lesion context. Each used a hierarchical classifier head producing three coupled outputs: diagnostic group (benign vs malignant), major category (melanoma vs non-melanoma skin cancer), and specific diagnosis constrained to a clinically plausible path (for example, malignant → melanoma → nodular melanoma). The secondary model superseded the primary model where appropriate. Calibrated probabilities were mapped to five triage categories (likely malignant; likely malignant – SCCiS; pre-malignant actinic keratosis; likely benign; and no result/poor image), which were shown to the nurse. The primary model was trained on 137,940 dermoscopic lesions from 46,195 patients, and the secondary model on 33,086 lesions enriched for AK and SCCiS cases. Both were evaluated on a held-out validation set of 34,386 lesions from 11,548 patients, stratified by diagnostic group, subtype, and anatomical site, with no patient or lesion shared with the training data. No lesion, image, or patient in the present analytic cohort appeared in the training or validation datasets. All training and validation labels were dermatologist consensus diagnoses. Post-hoc probability calibration used temperature scaling applied independently at each level of the hierarchy, with monotonicity constraints preventing logically inconsistent predictions. Triage thresholds were selected by Youden-index optimisation on the validation set, prioritising sensitivity for malignant and pre-malignant categories. The system functioned only as decision support and did not generate diagnoses or referral decisions.

### Clinical implementation

Lesions were imaged and triaged by registered nurses (or melanographers) trained specifically in dermoscopic skin-cancer screening. All nurses completed the same structured training programme before independent practice (theoretical modules in lesion selection, dermoscopy, and imaging standardisation; supervised in-clinic practice; competency assessment; internal and externally accredited certification), with ongoing competency audits and continuing professional development **(Supplementary Note 1)**. This training was identical across the AI-assisted and standard arms. However, nurses in AI-assisted clinics additionally received training in the use of the AI decision-support tool. The nurse-experience tiers used in this analysis therefore reflect post-certification experience among nurses who had all completed this common pathway. Nurses performed a full skin examination and captured clinical and dermoscopic images. In AI-assisted clinics, the AI processed each dermoscopic image at the point of capture and displayed a malignancy-risk score and risk band to the nurse on the clinical workstation. The nurse decided which lesions to forward for teledermatology assessment, with discretion to incorporate or disregard the AI output.

The AI did not forward lesions or determine any subsequent clinical action. In standard clinics, no AI output was shown. In both pathways, the teledermatologist reviewed the forwarded images and metadata and recorded a diagnosis and a recommended clinical action (no action, safety-netting, or intervention). Because lesion selection was performed by the nurse, the analysed lesions are those forwarded for assessment rather than every lesion imaged. This introduces potential selection bias, which is addressed in the Limitations.

### Statistical analysis

Lesions were the unit of analysis, following previous large real-world teledermoscopy and dermatology-AI evaluations [23, 24]. To account for clustering of lesions within patients and clinics, we used cluster-robust standard errors and within-clinic sensitivity analyses. Patient-and visit-level analyses are reported in the **Supplementary Notes 2** and **3** respectively. The primary analysis was a logistic regression of malignant status on AI adoption, adjusted for nurse experience tier. The AI odds ratio from this model is the primary effect estimate. We assessed whether the AI association varied with nurse experience by adding an AI-by-tier interaction and testing it with a likelihood-ratio test, and we report tier-specific estimates. We ran four pre-specified sensitivity analyses: adding Fitzpatrick phototype to the primary model, a within-clinic fixed-effects model that uses only within-site variation in AI use and removes fixed differences between clinics and between countries, and cluster-robust generalised estimating equations with an exchangeable working correlation, clustered by clinic and separately by patient. Analyses used Python 3.12 (statsmodels for regression, scipy.stats for descriptive comparisons). Odds ratios are reported with 95% confidence intervals, and the primary outcome was tested at a two-sided alpha of 0.05.

### Data availability and reporting

De-identified data are stored in a secure research environment and may be shared under material transfer agreements. The AI model is proprietary to MoleMap. Trained model weights cannot be released, but the architecture and preprocessing are described above. Reporting follows STROBE [25] guidance (**Supplementary Checklist**).

## RESULTS

### Study cohort

Nurses assessed 1,625,919 lesions across the clinics during the study period. Of these, 1,126,246 received a completed teledermatologist diagnosis and formed the basis for all rates. The remaining 499,673 were assessed by the AI but not individually diagnosed. As they were recorded in the study data for AI-assisted visits only, they were excluded from rate denominators. After removing non-screening services (22,619 lesions) and Australian AI-assisted visits (1,245 lesions), the analytic cohort comprised 1,102,382 lesions from 98,422 patients across 111,299 visits at 577 sites (**Figure 1**). Standard screening accounted for 60,853 visits (54.7%) and 580,472 lesions (52.7%), of which 198,005 (34.1%) were in New Zealand and 382,467 (65.9%) in Australia. AI-assisted screening accounted for 50,446 visits (45.3%) and 521,910 lesions (47.3%), all from New Zealand. The standard arm drew on 545 sites and the AI-assisted arm on 193. Across the cohort, 22,422 lesions were diagnosed malignant.

### Baseline characteristics

The two arms were similar in sex (57.2% vs 58.0% female among visits with recorded sex; standardised mean difference [SMD] -0.02) and in extreme Fitzpatrick scale (type I vs types V and VI). The arms differed in mean age (52.2 vs 47.2 years; SMD +0.33), mid-range phototypes (types II– IV; absolute SMD 0.10–0.14), and most in nurse experience (Tier I: 59.2% vs 45.4%; SMD +0.28) (**Table 1**). These differences are concentrated in the Australian component of the standard arm, reflecting the geographic imbalance in AI deployment (AI-assisted screening was available only in New Zealand). They do not account for the findings below. The within-clinic fixed-effects analysis, which removes any fixed difference between clinics and between countries, and the New Zealand-only analysis both retain the association (see Sensitivity analyses). Nurse experience is adjusted for directly in the primary model. Standardised mean differences (SMDs) are reported as descriptive measures of between-arm balance, with an absolute value above 0.10 (|SMD| > 0.10) taken to indicate a non-trivial imbalance. Confidence intervals are not reported because formal tests at this sample size would flag negligible differences. SMDs are not computed for “not recorded” categories, which reflect data availability rather than patient characteristics.

**Table 1.** Baseline characteristics and key outcomes by screening pathway. Percentages are within-arm. SMD = standardised mean difference (absolute SMD > 0.10 denotes a notable imbalance); SMDs are shown for baseline characteristics only. BCC, basal cell carcinoma; SCC, squamous cell carcinoma.

| Characteristic | Standard | AI-assisted | SMD |
| --- | --- | --- | --- |
| Visits, n (%) | 60,853 (54.7) | 50,446 (45.3) | – |
| Lesions, n (%) | 580,472 (52.7) | 521,910 (47.3) | – |
| Mean age, years | 47.2 | 52.2 | +0.33 |
| ***Sex (% of visits with recorded sex)*** |  |  |  |
| Female | 58.0 | 57.2 | -0.02 |
| Male | 42.0 | 42.8 | +0.02 |
| Sex not recorded (% of all visits) | 19.8 | 22.7 | – |
| ***Fitzpatrick phototype (% of visits)*** |  |  |  |
| Type I | 13.5 | 13.7 | +0.01 |
| Type II | 49.5 | 56.5 | +0.14 |
| Type III | 30.8 | 26.1 | -0.10 |
| Type IV | 4.5 | 2.5 | -0.11 |
| Types V and VI | 0.3 | 0.1 | -0.04 |
| Not recorded | 1.5 | 1.1 | – |
| ***Nurse experience tier (% of visits)*** |  |  |  |
| Tier I (0–2 years) | 45.4 | 59.2 | +0.28 |
| Tier II (2–4 years) | 21.6 | 14.0 | -0.20 |
| Tier III (>4 y) | 33.0 | 26.8 | -0.14 |
| ***Outcomes (per 1,000 lesions)*** |  |  |  |
| Malignancy detection rate | 15.7 | 25.5 | – |
| BCC detection rate | 7.32 | 14.20 | – |
| SCC detection rate | 3.37 | 6.35 | – |
| Melanoma detection rate | 0.82 | 1.81 | – |
| No-action recommendations | 866 | 832 | – |
| Safety-netting recommendations | 88 | 101 | – |
| Intervention recommendations | 45 | 67 | – |
| Biopsy/excision recommendations | 16.16 | 20.93 | – |
| Number needed to biopsy | 1.03 | 0.82 | – |

### AI diagnostic performance

The AI system was evaluated in two stages against a dermatologist reference standard. An initial validation on the held-out development set, and a subsequent production-grade re-validation on an independent dataset later from the study period. On the held-out development validation set (34,386 lesions from 11,548 patients, stratified by diagnostic group, subtype, and site), the primary model discriminated malignant from benign lesions with an AUROC of 94.2%, at 94.3% sensitivity and 70% specificity. Melanoma discrimination reached an AUROC of 87.1%, at 96.6% sensitivity and 4.9% specificity and the secondary model distinguished SCCiS from AK with an AUROC of 91%, at 84% sensitivity and 82% specificity. The system was then re-validated at production grade on 252,229 dermoscopic images (18,295 patients; February–April 2025) from the present analytic cohort, independent of the development dataset. Benign-versus-malignant discrimination reached an AUROC of 0.955. At the deployed operating point, the model identified 94% of reference-standard malignancies (sensitivity 94% and specificity 81%). Consistent with a design that prioritises sensitivity, 30% of its malignant calls were positive on the reference standard, the remainder passing to teledermatologist review. Melanoma discrimination reached an AUROC of 0.936 (sensitivity 98%) but at low precision, so most melanoma-level flags were not melanoma. These false positives are resolved downstream by the teledermatologist, who assesses every forwarded lesion. Within intraepithelial-carcinoma predictions, the secondary model distinguished SCCiS from AK with an AUROC of 0.847 (sensitivity 73%, specificity 81%). Sensitivity was maintained across skin phototypes, but specificity declined on darker skin (Fitzpatrick IV-V) increasing false-positive flags. Full model details and dataset composition information are provided in **Supplementary Note 4.**

### Primary outcome – malignancy detection rate

The malignancy detection rate was 25.5 per 1,000 lesions with AI assistance and 15.7 per 1,000 without. Unadjusted, this gives an odds ratio of 1.63 (95% CI 1.59– 1.68). However, AI was deployed disproportionately at clinics staffed by less-experienced nurses, whose baseline detection rate is lower, which dilutes the unadjusted comparison. After adjustment for nurse experience tier, AI assistance increased the odds of malignancy diagnosis by 73% (adjusted OR 1.73, 95% CI 1.68–1.77; p < 0.001). This adjusted odds ratio is the study’s primary result. The unadjusted, interaction, subgroup, and sensitivity estimates that follow are reported to characterise its robustness and variation across nurse experience, not as alternative primary estimates.

### Effect across nurse experience

The association was present in every experience tier but was not uniform. An AI-by-tier interaction was significant (likelihood-ratio test, p < 0.001; null hypothesis: no interaction between AI use and nurse experience). Detection rose from 13.5 to 21.2 per 1,000 for Tier I nurses (OR 1.58, 95% CI 1.52–1.64), from 16.4 to 29.9 for Tier II (OR 1.85, 95% CI 1.74–1.97), and from 18.8 to 34.4 for Tier III (OR 1.87, 95% CI 1.78–1.95) (**Figure 2**). The absolute gain was therefore largest for the most experienced nurses.

**Figure 2.**
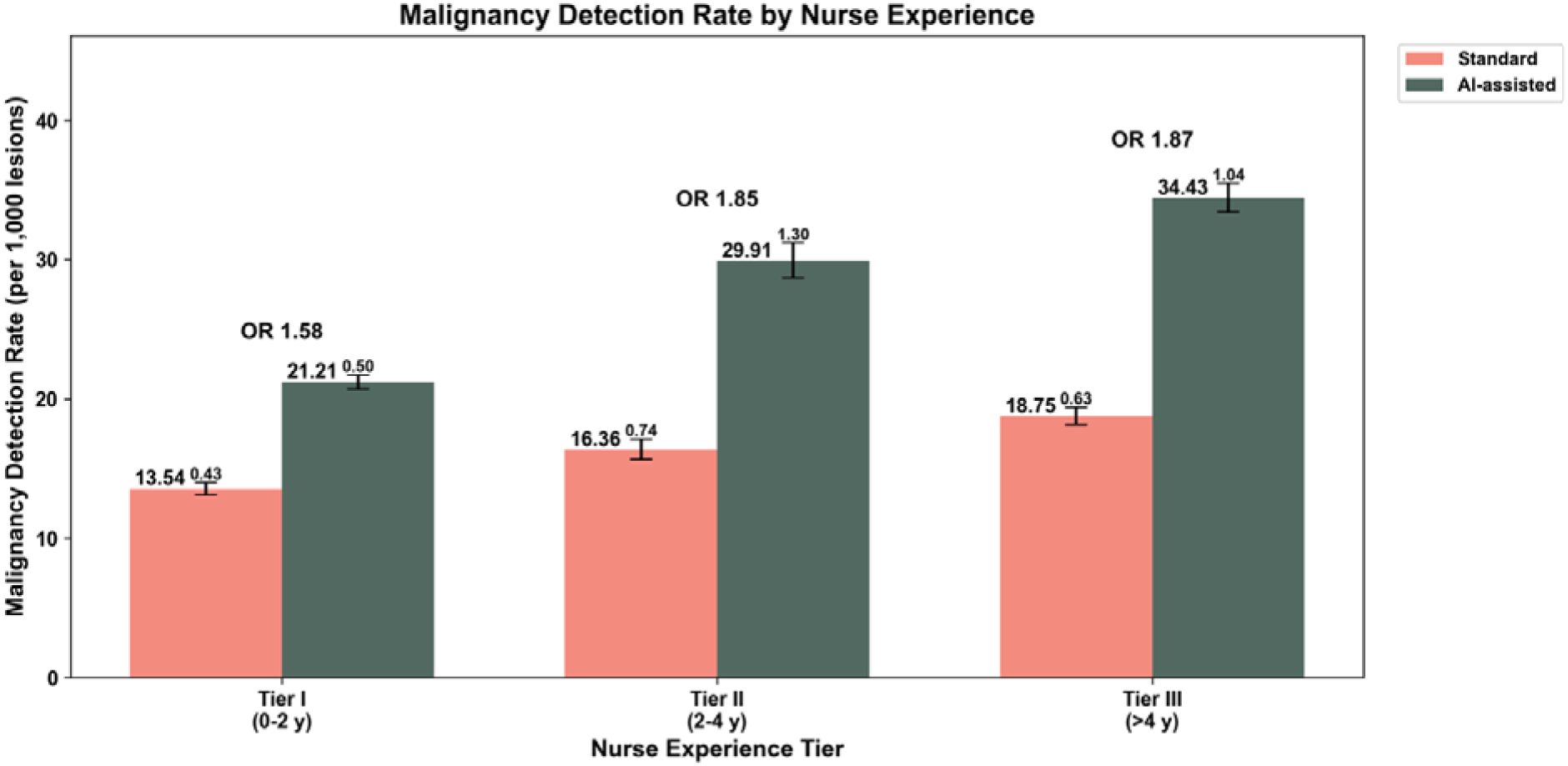
Malignancy detection rate by nurse experience tier. Detection rate (teledermatologist-diagnosed malignancies per 1,000 lesions assessed) for nurses stratified into three experience tiers. Bars represent observed rates; error bars are 95% Wilson confidence intervals. Green bars, with AI decision support; coral bars, without AI. AI-assisted screening was associated with higher detection in all three tiers: Tier I (0–2 years), 13.5 to 21.2 per 1,000 (OR 1.58; 95% CI 1.52–1.64); Tier II (2–4 years), 16.4 to 29.9 (OR 1.85; 95% CI 1.74–1.97); Tier III (>4 years), 18.8 to 34.4 (OR 1.87; 95% CI 1.78–1.95). The AI × experience interaction was significant (p < 0.001). OR, odds ratio; CI, confidence interval.

### Sensitivity analyses

The primary estimate was stable across pre-specified sensitivity analyses. Adding Fitzpatrick phototype left it essentially unchanged (OR 1.73, 95% CI 1.68–1.78). Restricting the comparison to within-clinic variation in AI use, which removes all fixed differences between clinics and between countries, did not weaken the association but slightly strengthened it (OR 1.83, 95% CI 1.73–1.93), indicating that the pooling of Australian and New Zealand standard-care visits does not drive the result. Restricting to New Zealand alone gave a more conservative estimate (adjusted OR 1.63, 95% CI 1.57–1.70). Accounting for the non-independence of lesions with cluster-robust generalised estimating equations attenuated the point estimate, as expected for a population-averaged model, while remaining clearly significant. Clinic-level clustering produced a more conservative estimate (OR 1.25, 95% CI 1.20–1.31) than patient-level clustering (OR 1.39, 95% CI 1.34–1.44), reflecting greater correlation of lesions within clinics than within patients. Visit- and patient-level analyses are reported in the **Supplementary Note 5**.

### Detection by Malignant Subtype

AI assistance was associated with higher detection of all three major subtypes: basal cell carcinoma (n = 11,657) rose from 7.32 to 14.20 per 1,000 (OR 1.95, 95% CI 1.88–2.03), squamous cell carcinoma (n = 5,272) from 3.37 to 6.35 (OR 1.89, 95% CI 1.79–2.00), and melanoma (n = 1,422) from 0.82 to 1.81 (OR 2.21, 95% CI 1.98–2.46) (**Figure 3**). The larger relative estimate for melanoma is influenced by the lower melanoma detection rate in the Australian standard-care component. Within-country comparison is reported in the **Supplementary Note 5.**

**Figure 3.**
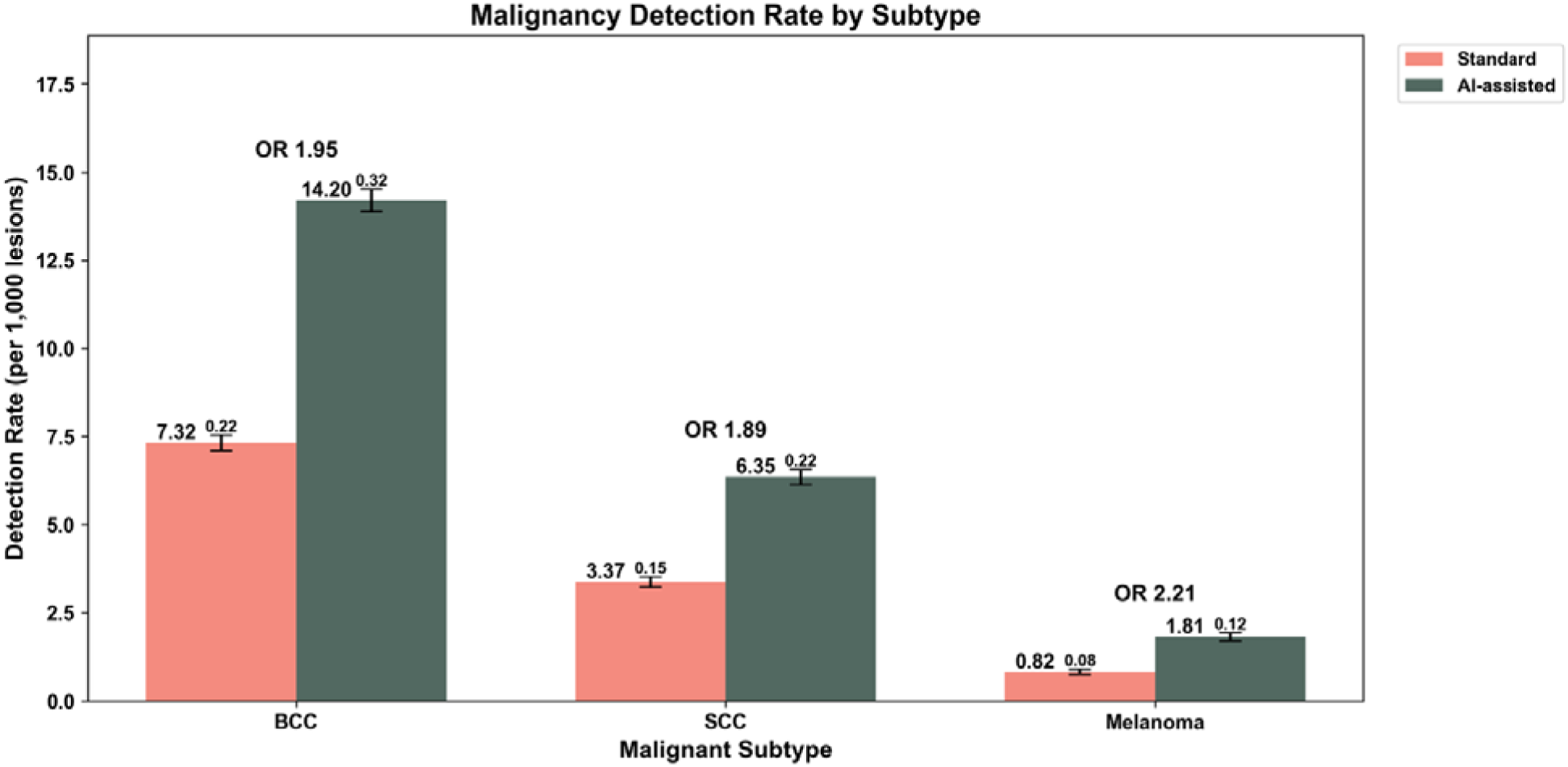
Malignancy detection rate by subtype. Bars denote screening pathway: green, with AI decision support; coral, without AI. Subtype-stratified detection rate for the three major malignant subtypes, with and without AI decision support. BCC, 7.32 to 14.20 per 1,000 (OR 1.95; 95% CI 1.88–2.03); SCC, 3.37 to 6.35 per 1,000 (OR 1.89; 95% CI 1.79–2.00); melanoma, 0.82 to 1.81 per 1,000 (OR 2.21; 95% CI 1.98–2.46). Subtype labels are teledermatologist diagnoses; no histopathological confirmation was available. The melanoma comparison is influenced by the lower melanoma detection rate in the Australian standard-care component (**Discussion**). BCC, basal cell carcinoma; SCC, squamous cell carcinoma; OR, odds ratio; CI, confidence interval.

### Clinical Action Recommendations

AI assistance shifted recommended management away from inaction and towards active investigation. In absolute terms, AI-assisted screening was associated with 21 additional intervention recommendations and 13 additional safety-netting recommendations per 1,000 lesions, offset by 34 fewer no-action recommendations (**Figure 4**). As rates, no-action recommendations fell from 866 to 832 per 1,000 (OR 0.77, 95% CI 0.76–0.77), safety-netting rose from 88 to 101 (OR 1.16, 95% CI 1.15–1.18), and intervention (biopsy, excision, or superficial treatment) rose from 45 to 67 (OR 1.51, 95% CI 1.48–1.53). All three action categories differed significantly between arms (p < 0.001 for each).

**Figure 4.**
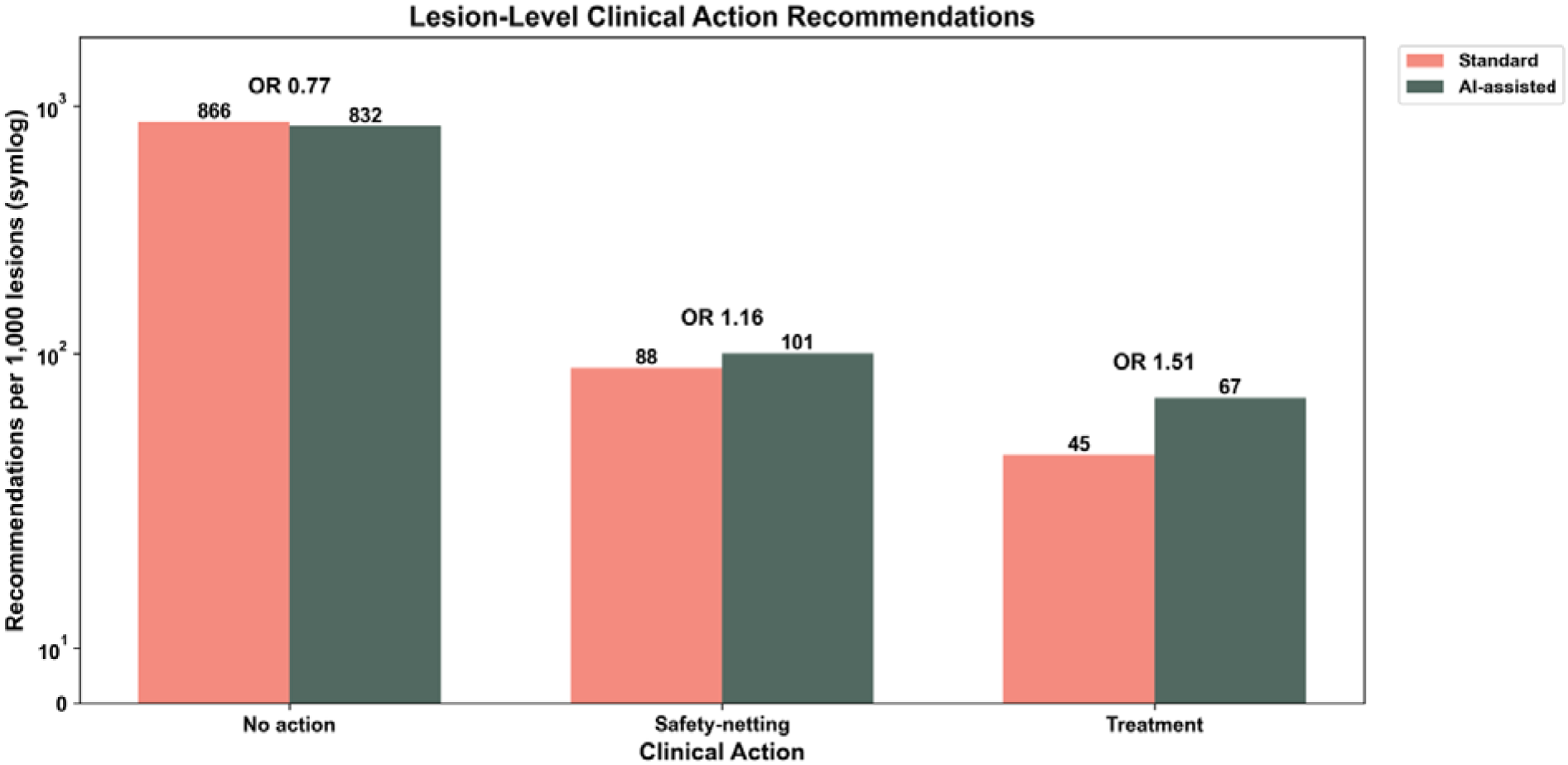
Lesion-level clinical action recommendations. Distribution of teledermatologist-recommended clinical actions across three categories. Bars denote screening pathway: green, with AI decision support; coral, without AI. AI-assisted screening was associated with a shift away from no-action recommendations (866 to 832 per 1,000; OR 0.77) and toward safety-netting (88 to 101 per 1,000; OR 1.16) and treatment (45 to 67 per 1,000; OR 1.51); all p < 0.001. The y-axis uses a symmetric-log (symlog) scale to display the large no-action category alongside the smaller categories. OR, odds ratio.

### Biopsy recommendations relative to malignancies detected

Biopsy or excision was recommended for 16.16 per 1,000 lesions under standard screening and 20.93 per 1,000 with AI. Expressed against the malignancies detected, this corresponds to 1.03 biopsy-or-excision recommendations per diagnosed malignancy under standard screening and 0.82 with AI. The rise in recommended procedures was more than matched by the rise in malignancies detected, indicating improved efficiency: fewer unnecessary procedures per malignancy identified. These figures describe recommendations recorded by the teledermatologist, not procedures performed, and no histopathology was available to confirm them.

### Safety

No adverse events attributable to screening or to AI-supported decisions were recorded during the study period. However, the dataset extract did not include systematic prospective adverse-event capture.

## DISCUSSION

This study evaluated whether embedding an AI decision-support tool into routine, nurse-led skin cancer screening changes malignancy detection and clinical management. Across more than 1.1 million lesions, AI-assisted screening was associated with a higher malignancy detection rate, and the difference held across every nurse-experience tier and all three major malignant subtypes. The most likely explanation is that AI changes which lesions nurses forward, though alternative mechanisms cannot be excluded. In the AI-assisted arm, the nurse saw a malignancy-risk score at the point of image capture, so the association plausibly reflects altered lesion selection *i.e.,* more malignant lesions sent on for diagnosis, and fewer clearly benign ones. This interpretation is consistent with the shift in recommended management: more intervention and safety-netting recommendations and fewer no-action recommendations, without a disproportionate rise in biopsy requests. However, a possible change in teledermatologist behaviour cannot be separated from these data.

While AI assistance was associated with higher detection across all experience tiers, the absolute gain was larger for more experienced nurses. One explanation is that experience and AI are complementary. An experienced nurse may be better placed to act on a risk score, forwarding a lesion she might otherwise have deprioritised while still setting aside clearly benign ones. A less experienced nurse may lack the confidence to override the AI output or may over-rely on it. Differences in case mix or in how AI was taken up at different clinics cannot be excluded. Regardless of the mechanism, the finding argues against treating AI as a substitute for experience. It raised detection most where clinical judgement was strongest. This aligns with human-factors research emphasising the importance of integrating AI design with clinical workflows and user expertise [26].

AI assistance was also associated with higher detection of melanoma, basal cell carcinoma, and squamous cell carcinoma. The teledermatologist diagnosis was the reference standard throughout, so these are gains in dermatologist-diagnosed malignancy rather than histologically confirmed disease. The larger relative estimate for melanoma is partly an artefact of lower melanoma detection in the Australian standard-care component and should not be over-interpreted.

### Implementation and regulation

In New Zealand, software as a medical device that carries a diagnostic claim requires registration, and privacy and security obligations are legislated, but constraints on performance claims are currently lighter than in Australia and many other jurisdictions *i.e.,* fewer requirements for independent clinical validation before deployment. In the workflow evaluated here, the teledermatologist retains responsibility for every diagnosis and management decision, with the nurse retaining discretion over lesion selection. This division of responsibility is an important safeguard when a decision-support tool is deployed at scale, as it prevents autonomous decision-making and ensures human oversight of all clinical decisions.

### Future work

Three directions matter most. First, and most important, prospective linkage to histopathology for all recommended procedures would let the teledermatologist reference standard itself be validated. The absence of pathology is the central evidential gap, as it prevents confirmation that improved dermatologist-diagnosed detection reflects true malignancies rather than diagnostic disagreement. Second, the equity of AI benefit across skin phototypes requires dedicated study. Pre-deployment testing showed specificity declining on darker skin, and this cohort was predominantly light-skinned, so we could not evaluate benefit in patients with Fitzpatrick V–VI skin. Two complementary directions follow. Prospective evaluation in more diverse or higher-incidence populations is needed to measure real-world benefit, though such cohorts are hard to assemble where skin cancer is uncommon in these groups. On the model side, the observed performance gradient may be reducible through training-side mitigations by reweighting or oversampling of under-represented phototypes, and, more tentatively, synthetic or augmented imagery to enrich scarce training examples though the fidelity of generated dermoscopic images for darker skin is itself unproven and would require validation against real cases before clinical use. Third, economic evaluation is needed to weigh earlier detection against added investigation and follow-up burden, though skin-cancer cost modelling is complex and rarely attempted. Health systems and payers would be the primary users of such analyses to inform implementation decisions.

## LIMITATIONS

The principal strengths of this study are its scale, comparable to or exceeding most real-world clinical AI evaluations in dermatology (1,102,382 lesions from 98,422 patients at 577 sites), its conduct within a single network under uniform protocols, and the within-clinic contrasts that allow between-site confounding to be removed. These are counterbalanced by several limitations that follow from the analysis being observational and from the data available to us.

### Non-randomised allocation and country

Clinics were not randomised to AI. Allocation followed deployment, producing imbalances in nurse experience, age, and phototype (Table 1). We adjusted for nurse experience and tested robustness through within-clinic and New Zealand-only analyses, but residual confounding from unmeasured variables (such as differences in patient case mix, clinic infrastructure, or AI uptake patterns) cannot be excluded. The AI-assisted arm is entirely from New Zealand while the standard arm pools New Zealand and Australian visits, so country is aligned with exposure. Because Australian standard care has a lower detection rate than New Zealand standard care, pooling lowers the comparator and widens the association relative to the New Zealand-only estimate (adjusted OR 1.73 pooled versus 1.63 within New Zealand). The within-clinic estimate, which is unaffected by fixed country differences, supports the finding, and we treat the New Zealand-only estimate as the more conservative one.

### Reference standard and blinding

No histopathology was available in the study data, so the teledermatologist diagnosis is the sole reference and could not be checked against pathology. Teledermatologists were not formally blinded to AI outputs, which were visible in the clinical platform, so an influence on their diagnoses or recommendations cannot be ruled out. Individual reviewer thresholds were not modelled, so variation in diagnostic criteria across teledermatologists could not be assessed.

### Data provided and lesion selection

Several fields that MoleMap records were not provided in the study extract (such as lesion anatomical site which may affect malignancy risk and complete patient sex), so could not be analysed. Lesions assessed by the AI but not individually diagnosed were available only for AI-assisted visits, so the denominator of lesions examined is characterised asymmetrically between arms and these lesions were excluded from rates. Because lesion selection was performed by the nurse, the cohort comprises forwarded lesions rather than every lesion imaged. The observed association between AI and malignancy detection operates through two mechanisms: (1) changed lesion selection by nurses (forwarding more malignant lesions when shown AI risk scores) and (2) changed diagnosis by teledermatologists. These data cannot separate these mechanisms, so the relative contribution of each is unknown.

### Analytic structure

Lesions are clustered within patients and clinics, and the primary model treats them as independent. Cluster-robust and within-clinic analyses confirm the association persists; the point estimate is attenuated under the population-averaged (GEE) specification, as expected for a marginal model, but visit- and patient-level estimands, which are insensitive to the number of lesions examined per visit, are consistent with the lesion-level result (**Supplementary Note 5**).

### Generalisability

This study was conducted within a single clinical network (MoleMap) in two high-income countries with predominantly light-skinned populations. Whether findings generalise to other healthcare systems, AI algorithms, or more diverse populations remains unclear.

## CONCLUSION

In this real-world evaluation of 1,102,382 lesions from 98,422 patients screened across 577 sites, real-time AI decision support was associated with a higher malignancy detection rate. This is, to our knowledge, the largest real-world evaluation of AI decision support in a nurse-led screening network. AI likely improved malignancy detection by helping nurses forward lesions most likely to be malignant while reducing unnecessary referral of benign lesions, with improved efficiency of recommended biopsies (0.82 vs. 1.03 per diagnosed malignancy) and higher detection across all major subtypes. Absolute gains were larger among more experienced nurses, indicating that AI is most effective when integrated with clinical experience rather than as a substitute for it. These findings rest on a teledermatologist reference standard without histopathological verification. They should be interpreted as strong evidence of association rather than proof of causation. AI performance varied by skin phototype, with specificity declining on darker skin (Fitzpatrick IV–VI), warranting attention during implementation to ensure equitable outcomes. Whether improvements translate into earlier melanoma stage and improved survival remains the key question, requiring prospective linkage to histopathology and long-term outcome data. AI should be implemented as a decision-support tool that complements clinical judgment, not as a replacement for experienced nurses or teledermatologists, with prospective monitoring of clinical outcomes and attention to equity across all populations.

## Data Availability

All data produced in the present study are available upon reasonable request to the authors

## Declarations

### Ethics

Approved by the University of Melbourne Human Research Ethics Committee (HREC ID 30352). The study used de-identified data from patients who had consented, under MoleMap’s standard data-use consent, to the use of their information and images for research.

### Author contributions

N.S. led the study and contributed to conceptualisation, data curation, formal analysis, investigation, methodology, visualisation, and writing of the original draft. N.D. contributed to methodology, data analysis and validation. K.W. and L.W. contributed to data curation, software development, resources, and project administration. A.D. contributed to methodology, statistical analysis, and validation. A.O. contributed to investigation, clinical validation, and resources (teledermatology reference standard). M.P. contributed to conceptualisation, funding acquisition, project administration, and supervision. J.V. contributed to conceptualisation, data curation, resources, software, validation and supervision. All authors contributed to writing, review and editing, and approved the final manuscript.

### Use of AI tools

During the preparation of this manuscript, the authors used SparkAI (the University of Melbourne’s secure internal generative AI environment) via the institutional enterprise portal to improve language, grammar, and structural flow. After using this service, the authors reviewed and edited the content as needed and take full responsibility for the final text and the overall integrity of the publication.

### Data availability

The data analysed in this study are de-identified clinical records from the MoleMap teledermoscopy network. These data are not publicly available because they are proprietary to MoleMap and were collected under patient data-use consent that does not permit open release. De-identified data may be made available from MoleMap for non-commercial research under a material transfer agreement, subject to ethics and privacy approvals. Requests should be directed to the corresponding author and will be referred to MoleMap. The AI model is proprietary to MoleMap. The trained model weights cannot be released, but its architecture and pre-processing are described in the Methods and Supplementary Information.

